# Neural Correlates of Cognitive Alterations and Minor and Structured Hallucinations in Parkinson’s Disease

**DOI:** 10.64898/2026.07.06.26357376

**Authors:** Lada Kohoutová, Jevita Potheegadoo, Léa Duong Phan Thanh, Sara Stampacchia, Marie E. Maradan-Gachet, Julien F. Bally, Cécile A. Hubsch, Mayte Castro Jimenez, Vanessa Fleury, Judit Horvath, Benoît Wicki, Paul Krack, Fosco Bernasconi, Olaf Blanke

**Author notes:** Please address corespondence to: Prof. Olaf Blanke, Laboratory of Cognitive Neuroscience, Bertarelli Foundation Chair in Cognitive Neuroprosthetics, Neuro-X Institute, Campus Biotech Chemin des Mines 9, 1202 Geneva Switzerland.

## Abstract

**Background:** Hallucinations, ranging from minor (MH) to structured, are a common non-motor symptom in Parkinson’s disease (PD). Structured hallucinations have been associated with altered functional connectivity (FC) between dorsal/ventral attention (DAN, VAN) and default mode (DMN) networks. As structured hallucinations are linked to rapid cognitive decline and MH are often viewed as their precursor, it is imperative to understand the neural basis of MH, and its relationship with cognitive alterations.

**Objectives:** We aimed to identify a whole-brain FC pattern associated with MH and alterations in attention-executive functioning in PD, leveraging a robotic procedure inducing presence hallucinations (riPH) experimentally, to which patients with hallucinations previously showed increased sensitivity.

**Methods:** Non-demented PD patients (*N* = 53) were categorized into three subgroups based on their hallucination symptoms: no hallucinations (nH; *n* = 19), MH (*n* = 18), and structured hallucinations, with or without MH (SMH; *n* = 16). We combined results from the riPH procedure and neuropsychological tests and applied multivariate methods capturing their shared variance in resting-state fMRI data across the three subgroups.

**Results:** We identified a distributed FC pattern more strongly expressed in patients with hallucinations (MH, SMH), and equally so across both groups, significantly associated with alterations in attention-executive functions and differences in riPH sensitivity. The pattern was primarily driven by FC between subcortical areas and visual network, DAN and DMN, and within-cerebellar and within-subcortical FC.

**Conclusions:** Our results highlight the role of subcortical-cortical connectivity in PD hallucinations, associated with cognitive alterations and already present in less advanced MH patients.

## Introduction

Parkinson’s disease (PD), although primarily characterized by motor symptoms including bradykinesia, rigidity and tremor, also manifests with debilitating non-motor symptoms such as hallucinations^1,2^. Hallucinations affect 30-60 % of PD patients^3–5^, and their complexity ranges from minor hallucinations (MH), which include visual illusions or misperceptions, passage and presence hallucinations (PH), to structured hallucinations, which are primarily visual, though auditory and tactile hallucinations can also occur^2,6^. Structured hallucinations generally appear in later disease stages^7–9^, and, crucially, have been consistently associated with faster cognitive decline and dementia^10–12^. MH occur earlier, sometimes even preceding motor symptoms^13^.

The neural systems underlying hallucinations in PD have been predominantly studied for structured visual hallucinations, which have been associated with altered visual processing, arguably based on the dominance of top-down over bottom-up processing^20,21,22,23^. Consistent with this, the attentional network hypothesis proposes that when PD patients process ambiguous visual stimuli, structured visual hallucinations may arise from decreased recruitment of the dorsal attention network (DAN), while overly relying on interpretation from the default mode (DMN) and ventral attention (VAN) networks, which couple with the visual network (VIS)^18,19^. Additionally, despite the subcortical origins of PD^20^, the functional role of subcortical areas in parkinsonian hallucinations remains less explored, with recent work associating alterations of the thalamus with hallucinations^21^. The neural mechanisms of MH are less well understood. Recent work reported alterations in functional connectivity (FC) within the DMN, alongside increased FC between the DMN and both the DAN and VIS in PD patients with MH^22^. However, divergent findings have been reported, specifically regarding no alterations in DMN-DAN^23^ and DMN-VIS^24^ coupling, and decreased FC between the DMN and DAN^24^.

Given that MH are viewed as a precursor to structured hallucinations^6,25^, which are associated with cognitive decline and dementia, understanding the relationship between MH and cognition is of particular interest. Indeed, MH have been suggested to carry an increased risk of cognitive decline^2,26^, but evidence remains mixed. While several studies failed to detect differences in cognitive performance between patients with MH and those without using standard neuropsychological tests^12,26^, others indicated a link between MH and impaired attentional control over interfering stimuli^27^, and more rapid decline of executive functions in longitudinal data^28^. These inconsistencies may reflect limitations of standard tools used to capture cognitive and neural correlates of hallucinations, whose investigation is inherently challenged by their transient-unpredictable and subjective nature^29^.

To address this, our group developed a robotic procedure that, via somatomotor stimulation, induces MH (specifically robot-induced PH; riPH) under controlled laboratory settings^30^. Critically, PD patients with MH exhibit heightened sensitivity to this riPH procedure compared to patients without^29,31^, and this sensitivity was associated with reduced frontal subcortical functions^31^. Moreover, this approach revealed altered FC between posterior middle temporal gyrus (pMTG) and inferior frontal gyrus (IFG), nodes of the DMN and VAN^29^, in PD patients with MH. Yet, the neural mechanisms linking hallucinations with cognitive deficits and riPH, and how they differ between MH and structured hallucinations, remain unexplored. Given that hallucinations in PD are recognized as a risk factor for cognitive decline, identifying these mechanisms is clinically relevant and may inform disease modifying therapies.

The current study integrates behavioral data from the riPH procedure with neuropsychological assessments and resting-state functional magnetic resonance imaging (rs-fMRI) from a group of PD patients (*N* = 53), categorized into three subgroups: patients with only MH (MH; *n* = 18), patients experiencing both MH and structured hallucinations, or only structured hallucinations (SMH; *n* = 16), and patients experiencing no hallucinations (nH; *n* = 19). We quantified whole-brain FC across cortical, subcortical, and cerebellar regions, and used partial least squares correlation (PLSC) to test multivariate brain-behavior relationships linking FC patterns with hallucination-related variables (including riPH sensitivity) and measures of attention-executive functioning.

## Methods

### Participants

Fifty-three patients with PD were included, diagnosed based on the Movement Disorder Society (MDS) criteria for PD^32^. These patients were a part of a larger multicentre project investigating hallucinations in PD. To assess occurrence and type of hallucinations, all patients completed a semi-structured interview with a neuropsychologist^31^, based on which they were classified into three subgroups: those who experienced at least one MH recurrently since disease onset and/or within the previous month (MH; *n* = 18), those who experienced at least one structured (auditory, olfactory, gustatory, tactile, and/or visual) hallucination, with or without MH, recurrently since disease onset and/or within the previous month (SMH; *n* = 16), and those who never experienced hallucinations (nH; *n* = 19). For socio-demographic and clinical details, including age, sex, education, disease duration, total levodopa equivalent daily dose (LEDD)^33^, dopamine agonist dose, cognitive impariment evaluated by Montreal Cognitive Assessment (MoCA)^34^, and evaluations of severity of non-motor and motor symptoms on the MDS Unified Parkinson’s Disease Rating Scale (MDS-UPDRS) and disease stage measured by the Hoehn and Yahr scale^35^ see **Table 1**. No patient had other neurological, psychiatric, or substance abuse disorders. All patients provided written informed consent prior to participation, approved by the Swiss cantonal ethics committees (protocol reference 2019-02275), in accordance with the Declaration of Helsinki.

### Robot task inducing presence hallucinations and neuropsychological evaluation

The riPH procedure was described previously^29–31,36^. In the current analysis, we categorized individuals as either “responders”, if their sensitivity to riPH increased with an increasing delay, or as “non-responders” otherwise. Group differences were tested with chi-square tests with false discovery rate (FDR) correction (see Supplementary methods). The neuropsychological evaluation was also described previously^31,37^. The main analysis of the current study uses the frontal subcortical score from the Parkinson’s Disease Cognitive Rating Scale (PD-CRS) reflecting attention-executive functioning^37^, but the posterior cortical PD-CRS score was also evaluated (see Supplementary methods).

### MRI data acquisition

Patients, in an on-medication state, underwent an MRI session in which a structural scan and 8 min of rs-fMRI were acquired using a 3 T Siemens Prisma scanner. High-resolution T1-weighted structural images were acquired using an MPRAGE sequence (TR = 2.2 s, TE = 2.96 ms, flip angle = 9°, base resolution = 256 × 256, voxel size = 1 mm isotropic). Functional images were obtained with a multiband accelerated echo-planar imaging (EPI) sequence (multiband factor = 8, TR = 700 ms, TE = 30 ms, flip angle = 52°, voxel size = 2 mm isotropic).

### fMRI data preprocessing

The fMRI data were preprocessed using SPM12 (https://www.fil.ion.ucl.ac.uk/spm/software/spm12/) and subsequently denoised using the CONN 22 toolbox (https://www.nitrc.org/projects/conn/). First, the functional images were realigned and corrected for distortion. Then, the T1-weighted structural image was coregistered to the mean realigned functional image. Next, the coregistered structural image was segmented to extract the gray matter, white (WM) matter, and cerebrospinal fluid (CSF) tissue masks. The images were then normalized into the Montréal Neurologic Institute (MNI) space and smoothed with a 6 mm Gaussian kernel. Finally, the functional data were denoised using motion parameters, volumes tagged at framewise displacement > 0.9 mm, and WM and CSF signal as regressors, and passed through a band-pass filter of a range from 0.008 Hz to 0.09 Hz. All subjects’ fMRI data passed a quality control check as the maximum percentage of motion-tagged volumes was 6.2 %.

### Estimation of FC

We parcellated each subject’s whole brain functional data using a combination of cortical region from the Schaefer atlas^38^ and subcortical and cerebellar regions from the Brainnetome atlas^39^, with periaqueductal grey and brainstem regions from previous studies^40,41^, resulting in 265 parcels. We then estimated FC matrices within each individual using Pearson’s correlation coefficient between each pair of mean time courses in brain parcels.

### Partial Least Square Correlation (PLSC) analysis

Given the multidimensional nature of hallucinations and cognition, we employed PLSC as implemented by the Matlab-based myPLS library (https://github.com/MIPLabCH/myPLS) to investigate brain-behavior relationship in a multivariate manner. Vectorized lower triangles of individual FC matrices were entered as the predictor variable, while subgroup assignment, binary riPH task data (responders vs. non-responders), and frontal subcortical PD-CRS scores constituted the outcome variable. To determine significant latent components (LCs), representing shared multivariate patterns of covariance between FC and all outcome variables, we ran permutation tests with 5,000 iterations. Then, within the significant LC, we performed a bootstrap test with 10,000 samples to identify edges significantly contributing to the given LC. Edge significance was evaluated by the bootstrap ratio (BSR) calculated as the observed salience divided by the standard deviation of the bootstrapped salience. Edges with BSR > 2 were considered significant as suggested by Krishnan et al.^42^. For interpretability, we used a threshold of top 1 % of BSR.

### Post hoc analysis of PLSC results

To further explore the significant LC, we analyzed the relationship between brain scores, that is, the dot product of the brain salience map and FC matrices, and outcome variables. For the frontal subcortical PD-CRS score, we computed Spearman’s *ρ* between brain scores and frontal subcortical scores within each subgroup. In the same way, we also assessed the contribution of each subscore of the frontal subcortical score. To evaluate differences in brain scores between riPH responders and non-responders and across subgroups, we used a two-way ANOVA with post hoc Tukey’s test. We also tested the association between the identified LC and variables of no interest, including age, education, disease duration, disease stage, dopamine agonists and total LEDD (see Supplementary methods).

## Results

### Clinical and neuropsychological evaluation

The three patient subgroups did not significantly differ in age, education, medication, disease duration or stage (**Table 1**). The frontal subcortical and posterior cortical PD-CRS scores were not significantly different between the subgroups (*F*(2) = 0.563, *p* = 0.573 and *F*(2) = 0.337, *p* = 0.716, respectively) (**Fig. 1a, b**).

**Figure 1.**
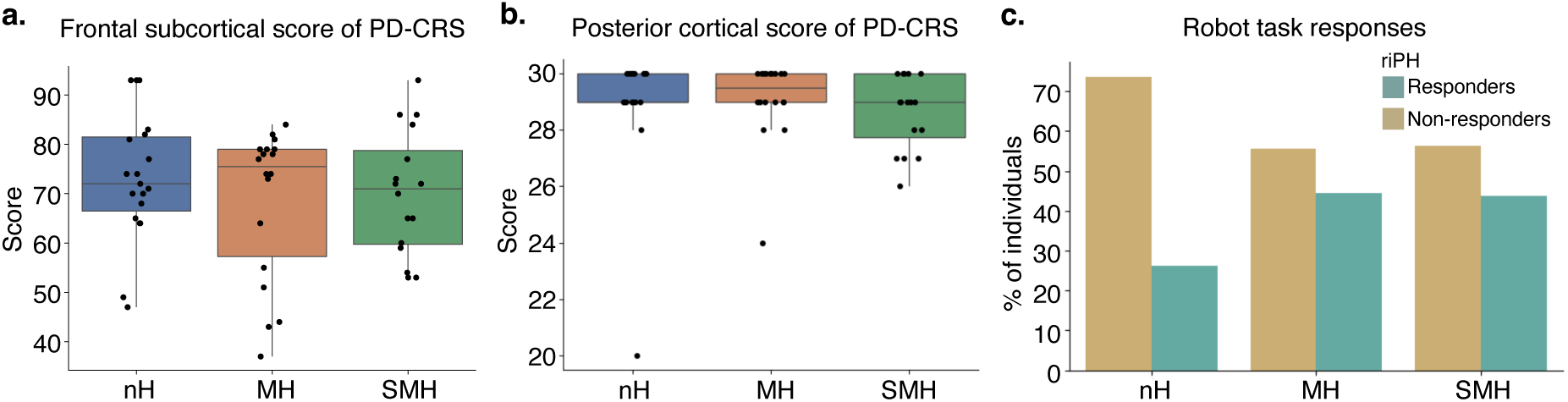
Cognitive scores measured by the PD-CRS and riPH responses. **a.** The frontal subcortical and **b.** the posterior cortical scores of the PD-CRS did not significantly differ between the patient subgroups. **c.** The bar plot shows, in each subgroup, the percentage of individuals who tended to report a feeling of presence with an increasing delay vs. those who did not. The nH subgroup showed a trend towards a larger percentage of non-responders (*p* = 0.039) which was not significant after an FDR correction (*p*_FDR_ = 0.117).

In MDS-UPDRS assessment, only MDS-UPDRS I, evaluating non-motor experiences of daily living, differed significantly between subgroups (*p* = 0.004), with both MH and SMH subgroups scoring significantly higher than the nH subgroup (*p* = 0.022 and *p* = 0.007, respectively). MH and SMH did not significantly differ in MDS-UPDRS I.

### Sensitivity to the riPH procedure

Before FDR correction, the nH subgroup showed significantly more “non-responders” than “responders” (χ²(1) = 4.263, *p* = 0.039), though this did not survive FDR correction (*pFDR* = 0.117) (**Fig. 1c**). MH and SMH did not differ significantly. A closer examination of the data with a linear model fitted to mean responses in each delay revealed a previously observed effect of delay (*t*(5) = 2.004, *p* = 0.047) without subgroup differences (**Supplementary Fig. 1**). However, the current data are a subset of data used in a previous study^31^ showing heightened sensitivity to the riPH procedure in patients with MH compared to those without, suggesting limited power given the smaller sample in the current study.

### PLSC analysis of rs-fMRI

The PLSC analysis revealed one significant LC (*pFDR* = 0.04) (**Supplementary Fig. 2a**), explaining 20.6 % of covariance, with behavior-brain scores correlation of *r* = 0.78, *p* = 5 ×10^-12^. The LC captured a robust multivariate association between an FC pattern and several components of the outcome variable (**Fig. 2a**). Specifically, the LC was significantly associated with the contrast of the two hallucination subgroups (MH and SMH) combined against nH. In interaction with this contrast, both frontal subcortical PD-CRS scores and riPH sensitivity (responses to the riPH procedure) showed significant correlations with the LC. The corresponding FC pattern was distributed across the whole brain. For interpretability, **Fig. 2b** shows top 1 % of loadings (BSR > 4.72) (all significant edges at BSR > 2 are shown in **Supplementary Fig. 2b**). **Fig. 2c** depicts the percentages of these loadings in each network pair, showing that FC between the subcortical areas (SBC, including the thalamus, caudate, putamen, and others) and VIS had the largest proportion of significant edges (8.8 %), suggesting that FC in this cortical-subcortical network pair was the most prominent contributor to the LC. The second largest proportion of significant features was found within SBC (3.6 %), followed by CRB (3.4 %), DAN and SBC (2.9 %), and SBC and DMN coupling (2.5 %). While all these SBC-centered edges were positively correlated with the behavioral outcomes, all significant edges within the CRB had negative loadings (see **Supplementary Fig. 3** for percentages of positive and negative loadings of significant edges in all network pairs). We performed the same PLSC analysis using the posterior cortical scores of the PD-CRS instead of the frontal subcortical scores, finding no significant LC (**Supplementary Fig. 4**).

**Figure 2.**
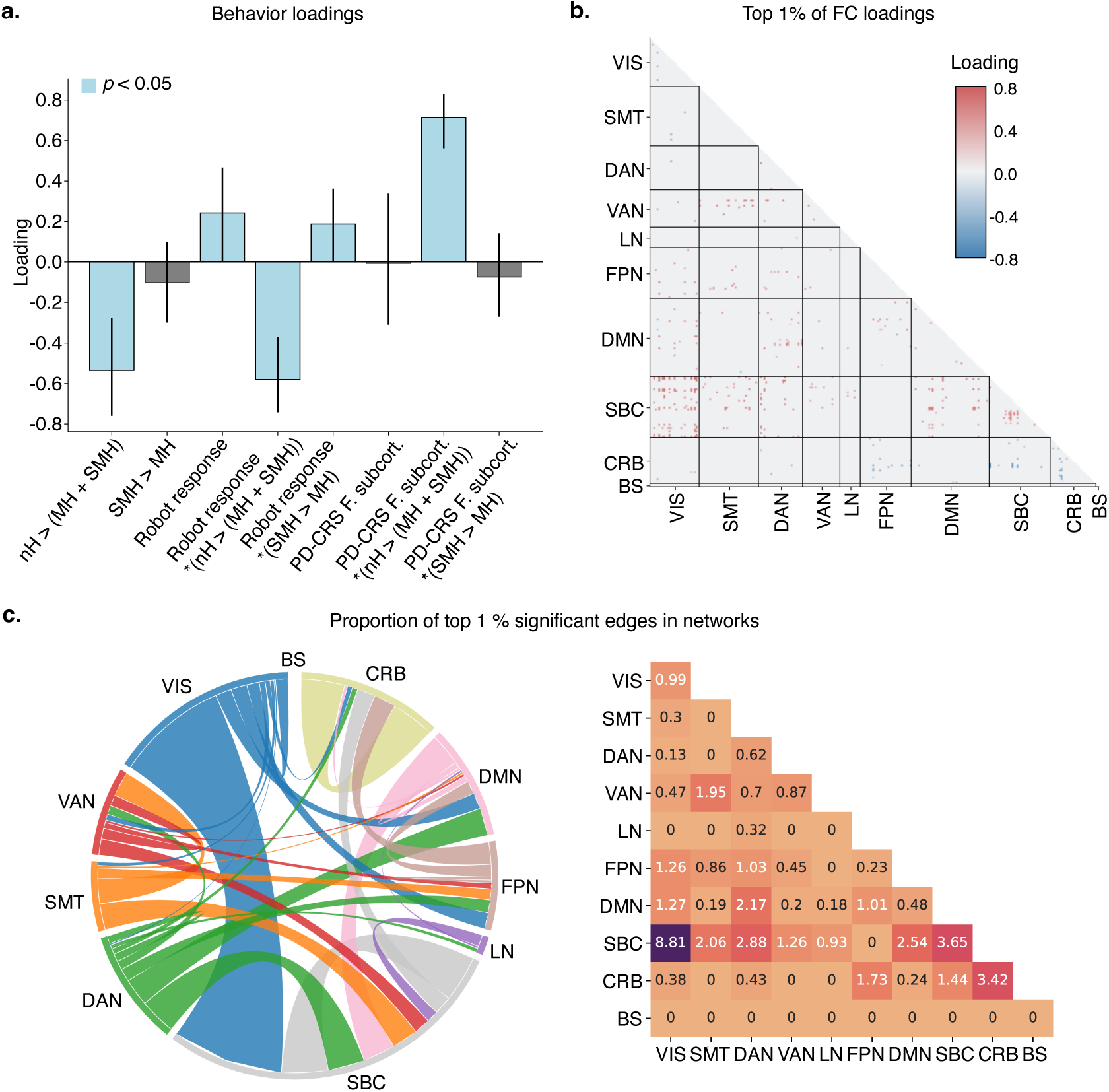
Loadings of the significant LC. **a.** The bar plot indicates loadings of all outcome variables. Significant variables are displayed in blue. **b.** The matrix shows top 1 % of loadings of the FC (the predictor variable) based on the BSR. **c.** The chord diagram and matrix summarize the percentage of the top 1 % of significant edges in each network. VIS – visual network; SMT – somatomotor network; DAN – dorsal attention network; VAN – ventral attention network; LN – limbic network; FPN – frontoparietal network; DMN – default mode network; SBC – subcortical network; CRB – cerebellum; BS – brainstem

### Link to neuropsychology and riPH

To closely examine the brain-behavior relationship in the significant LC, we first calculated the correlations between brain scores and frontal subcortical PD-CRS scores (**Fig. 3a**). All subgroups showed significant, but opposing relationships: positive correlations in MH and SMH subgroups (*ρ* = 0.587, *p*FDR = 0.016, and *ρ* = 0.5, *p*FDR = 0.049, respectively), and a negative correlation in the nH subgroup (*ρ* = -0.571, *p*FDR = 0.016). Details on which subtests composing the frontal subcortical score contributed to these effects can be found in **Supplementary Fig. 5.**

**Figure 3.**
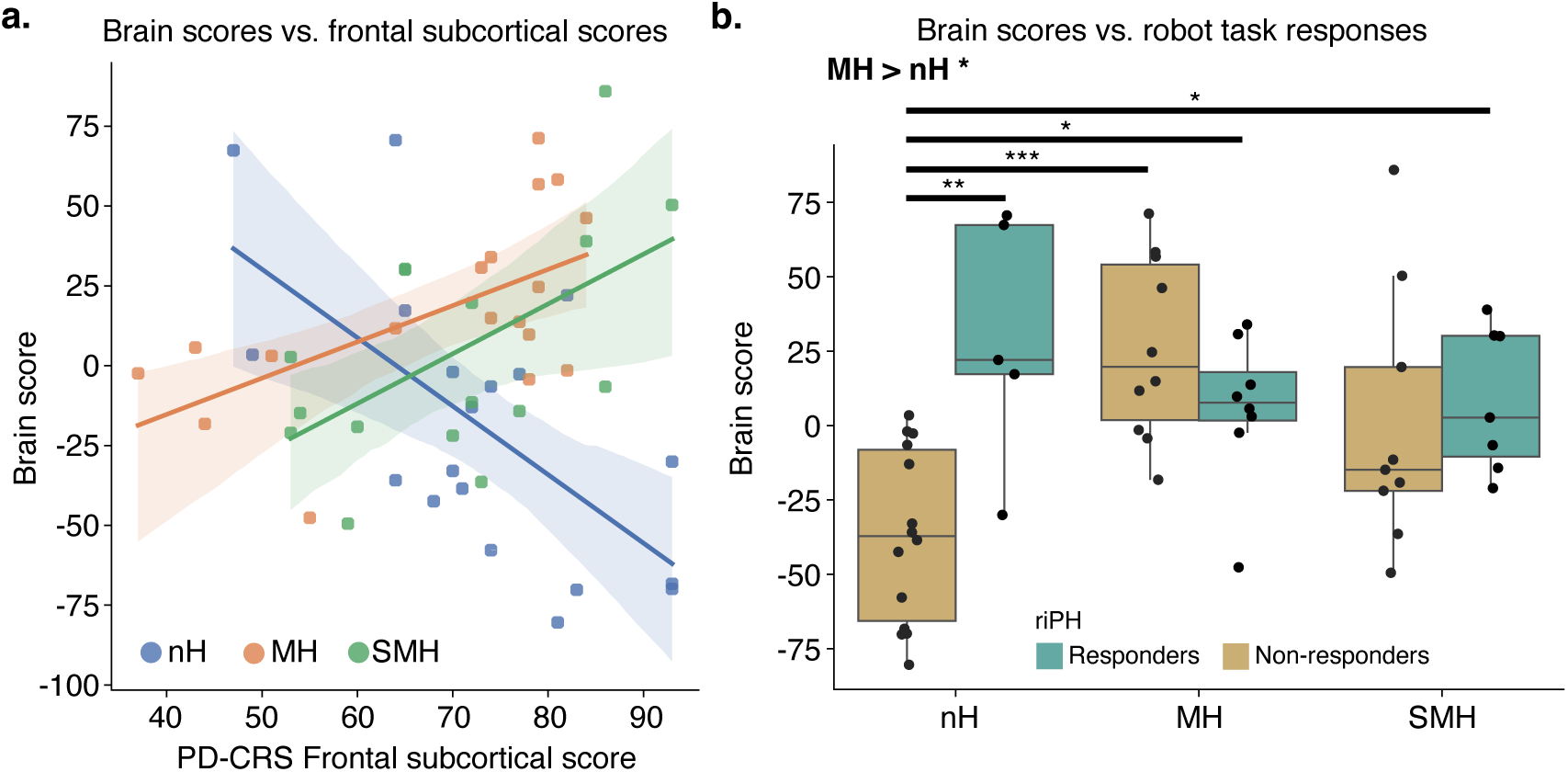
Post hoc analysis of the significant LC. **a.** The regression plot displays the relationship between the brain score and the frontal subcortical score, indicating that there was a positive relationship in the MH and SMH subgroups (*ρ* = 0.587, *p*_FDR_ = 0.016, and *ρ* = 0.5, *p*_FDR_ = 0.049, respectively), and a negative relationship in the nH subgroup (*ρ* = -0.571, *p*_FDR_ = 0.016). **b.** The boxplot shows differences between the three patient subgroups and riPH responder category. Except non-responders in the SMH subgroup, all other groups achieved higher brain scores than the non-responders in the nH subgroup.

A two-way ANOVA comparing the brain scores between subgroups and responses riPH procedure responses revealed a significant effect of subgroup (*F*(2) = 4.93, *p* = 0.011) and a subgroup × riPH response interaction (F(2) = 7.29, *p* = 0.002). Tukey’s post hoc test showed that MH had overall significantly higher brain scores than nH (mean difference = 36.49, *p* = 0.011). Regarding the patient subgroup × riPH response interaction, several interaction pairs achieved significantly higher brain scores than nH non-responders: MH responders (mean difference = 42.79, *p* = 0.0497), MH non-responders (mean difference = 62.91, *p* = 0.0003), and SMH responders (mean difference = 45.52, *p* = 0.0434). Critically, nH responders (i.e., those nH patients in whom riPH could be evoked using the the robotic procedure) did not significantly differ from either hallucination subgroup and reached significantly higher brain scores than nH non-responders (mean difference = 66.4, *p* = 0.0036; **Fig. 3b**). No significant differences were found between the MH and SMH subgroups.

Taken together, the identified FC pattern was more strongly expressed in subgroups with hallucinations, with opposing relationship with the frontal subcortical scores across subgroups. Furthermore, nH responders showed comparable levels of pattern expression to those observed in both MH and SMH, suggesting that patients without hallucinations in daily life who are sensitive to the riPH procedure and have low frontal subcortical cognitive scores show a similar degree of alignment to the FC pattern as patients with hallucinations (MH, SMH).

### Effects of nuisance variables

We found no significant relationships with nuisance variables, except in total LEDD that was significantly correlated with brain scores in the MH subgroup (*ρ* = -0.659, *p*FDR = 0.009) (**Supplementary Fig. 6**). However, after regressing out total LEDD from FC matrices and repeating the PLSC analysis, the originally identified LC pattern was largely preserved (see Supplementary results).

## Discussion

To investigate neural correlates of alterations in attention-executive functioning and their association with riPH across three subgroups of PD patients, we integrated whole-brain rs-fMRI data with neuropsychological measures and behavioral data from the riPH procedure. We identified a whole-brain cortical-subcortical multivariate FC pattern that (1) captured covariance of hallucinations and cognitive alterations in PD patients, (2) was more strongly expressed in hallucination subgroups, and (3) showed subgroup-dependent relationship with frontal subcortical (PD-CRS) task performance.

The identified significant LC was an FC pattern centered on subcortical structures (thalamus, basal ganglia). The largest proportion of significant edges was found in subcortical-visual network coupling. FC alterations in visual cortices have been often linked to structured visual hallucinations in PD^19,22,43,44^, and also to MH (as MH include visual illusions)^22,43^, consistent with reported decreased effective connectivity from the lateral geniculate nucleus (LGN) to visual cortex in PD patients with structured visual hallucinations^45^. Together with evidence that substantia nigra projects to the inferotemporal cortex via the thalamus^46,47^, this suggests an association between subcortical pathology and disrupted visual processing, potentially leading to visual hallucinations.

In the pattern, we also observed noteworthy contributions of SBC-DAN and SBC-DMN connectivity. Coupling between DAN and subcortical structures has been shown to be supported by subcortical nodes located in the caudate and the thalamus, including mediodorsal nuclei and pulvinar^48^. The pulvinar, together with the LGN, is implicated in visual processing and proposed as an integration hub linking the DAN and visual networks^49^, raising the possibility that altered thalamic visual relay and attentional hubs represent an important dysfunction in parkinsonian hallucinations. Mediodorsal thalamic nuclei have also been associated with DMN activation and its involvement in internally focused cognition^50^. SBC-DMN, as well as SBC-SBC coupling, has also been associated with longitudinal changes in executive functioning in PD^51^. Finally, our pattern exhibited minor contributions of SBC-VAN, and VAN connectivity has been linked to structured visual hallucinations in PD^18,24^. Taken together, while FC around the DAN and DMN has been frequently associated with structured visual, hallucinations in PD^18,22,24^, our findings suggest these cortical-subcortical alterations are closely tied to subcortically centered FC and, critically, also manifest in non-structured minor hallucinations.

Further testifying to the importance of basal ganglia and the thalamus in hallucinations in PD, we also found that FC within subcortical areas was strongly expressed in the identified pattern. Previous work primarily highlighted structural changes in the SBC, particularly a loss of the gray matter volume in striatal areas and thalamus in PD patients with structured visual hallucinations^52–54^, suggesting a structural basis that may be reflected in the altered FC, including connectivity with the visual cortex and DAN. Additionally, subcortical, and particularly thalamic nodes, have been found to be implicated in altered temporal functional dynamics underlying MH and structured visual hallucinations in PD^21^.

Another dominant contributor to the pattern, though rarely observed previously, was FC within the cerebellum. Previous work has linked visual hallucinations in PD to reduced cerebellar gray matter volume^55^. At the functional level, within-cerebellar FC has been shown to exhibit more subject-specific features in PD patients with MH compared to those without hallucinations and to relate to the frontal-subcortical PD-CRS scores^56^. Notably, in the current study, within-cerebellar FC contributed inversely to the LC pattern in contrast to the other discussed network pairs, indicating that FC within the cerebellum diverges from the dominant large-scale network configurations.

The identified multivariate pattern was differentially expressed across the patient subgroups, notably with stronger expression observed in hallucination subgroups relative to nH and with both hallucination subgroups showing positive associations with the frontal subcortical cognitive score. This indicates that weaker alignment with the pattern corresponded to worse cognitive performance in attentional and executive-frontal functions. Critically, this relationship was only observed in PD patients with hallucinations (MH, SMH), as this association was reversed and negative in nH patients. While previous work reported associations between FC and cognitive deficits specifically in patients with PD with MH^29,56^, our findings extend this by highlighting that this relationship is captured in a distributed, whole-brain multivariate connectivity pattern and differs from patients without hallucinations.

Our data showed no differences in the pattern expression between the MH and SMH subgroups. While some studies indicated that MH and structured visual hallucinations are driven by distinct mechanisms^57,58^, particularly in FC between the amygdala, insula, hippocampus, and fronto-temporal regions, others reported overlapping neural correlates^22,24^, specifically in within-DMN FC, and DMN’s coupling with visual areas and DAN. Patients with MH also showed structural changes in visual areas and the DMN^22^. In the present study, neural alterations in MH were comparable to those in SMH, including SBC-VIS, SBC-DAN and SBC-DMN contributions. Thus, our results align with the latter view, supporting the notion that hallucinations in PD form a single clinical-neural spectrum, rather than distinct neurobiological entities^6,25^. Notably, the two subgroups did not differ in any clinical and neuropsychological measure. We note that individuals from the SMH subgroup experienced not only structured hallucinations, but also MH, and the MH subgroup included patients experiencing simple visual illusions, indicating a substantial sensory modality overlap between the subgroups that may have contributed to their neural overlap. Together, this suggests that brain alterations between MH and SMH are minimal when tested in early-stage and mid-stages of PD.

The current study employs the riPH procedure, to which PD patients with hallucinations have shown higher sensitivity than those without^29,36,59^, and which has recently been shown to enhance detection of cognitive deficits in patients with MH^31^. Interestingly, nH individuals sensitive to riPH (i.e., in whom a hallucination-like state could be induced experimentally, despite no daily-life hallucinations) reached pattern expression levels comparable to MH and SMH. As these individuals also tended to have lower frontal subcortical cognitive scores, co-occurring riPH sensitivity and decreased cognitive performance appears linked to similar neural alterations even in patients without hallucination symptoms. These findings raise the possibility that such individuals may be at heightened risk of developing hallucinations in the future^31^, which could not be tested in the present sample. Combining riPH, with hallucination scoring and cognitive testing may help detect such patients before clinical hallucination onset and adapt potential treatments accordingly. Longitudinal studies are needed to investigate this possibility.

There are several limitations to this study. First, the current sample size is relatively small and lacking further validation data, which restricts statistical power and robustness of the results. Since the uniqueness of this clinical population naturally limits the sample size and practical constraints restrict the scan time, future studies could benefit from a deep sampling approach to increase statistical power. Second, as hallucinations in PD can develop and evolve over the course of the disease, longitudinal data are needed to explore progression and further elucidate whether our identified pattern captures early signs of worsening symptoms.

Furthermore, while the analysis used enables identification of whole brain patterns, it also possesses inherent challenges in interpretation of individual connections and regions.

In summary, our findings indicate that the relationship between hallucinations and deficits in attention-executive functioning in PD is reflected in a distributed FC pattern including subcortical and cerebellar regions, VIS, DAN and DMN, largely driven by cortical-subcortical connectivity. By integrating the riPH procedure, we further demonstrate that riPH sensitivity may reveal latent neural configurations even in patients without symptomatic hallucinations, potentially reflecting a risk of developing hallucinations later. Together, highlighting the value of multivariate whole-brain approaches to investigating neural correlates of complex phenomena in PD, the present results contribute to our understanding of systems-level mechanisms underlying hallucinations and cognitive deficits in PD, with potential implications for early risk identification and development of predictive models of symptom progression, enabling timely and targeted treatment.

## Data Sharing

Data sharing will require a formal data use agreement. Requests will be evaluated based on institutional and departmental policies to determine whether the data requested is subject to intellectual property or patient privacy obligations.

## Acknowledgement

The authors would like to thank all the patients for their valuable time and effort, in participating in the study.

Study data were collected and managed using REDCap electronic data capture tools hosted at the Swiss Federal School of Technology (Ecole Polytechnique Fédérale de Lausanne). REDCap (Research Electronic Data Capture) is a secure, web-based software platform designed to support data capture for research studies, providing 1) an intuitive interface for validated data capture; 2) audit trails for tracking data manipulation and export procedures; 3) automated export procedures for seamless data downloads to common statistical packages; and 4) procedures for data integration and interoperability with external sources.

## Author roles

(1) Research Project: A. Conception, B. Organization, C. Execution; (2) Statistical Analysis: A. Design, B. Execution, C. Review and Critique; (3) Manuscript Preparation: A. Writing of the First Draft, B. Review and Critique.

L.K.: 1A, 2A, 2B, 3A

J.P.: 1A, 1B, 1C, 3B

L.D.P.T.: 1B, 1C, 3B

S.S.: 2C, 3B

M.E.M-G.: 1B, 1C, 3B

J.F.B.: 1B, 3B

C.A.H.: 1B, 3B

M.C.J.: 1B, 3B

V.F.: 1B, 3B

J.H.: 1B, 3B

A. W.: 1B, 3B

P.K.: 1B, 3B

F.B.: 2C, 3B

O.B.: 1A, 3B

## Financial Disclosures of all authors (for the preceding 12 months)

This project was supported by two generous donors advised by CARIGEST SA, the first one wishing to remain anonymous and second one being *Fondazione Teofilo Rossi di Montelera e di Premuda*; the Bertarelli Foundation; Parkinson Schweiz, Leenaards foundation, Empiris foundation, Swiss National Science foundation (n° 320030_188798), Synapsis Foundation. OB is one of the inventors on patent US 10,286,555 B2 (Title: Robot-controlled induction of the feeling of a presence) held by the Swiss Federal Institute of Technology (EPFL) that covers the robot-induction of the presence hallucinations (riPH). O.B. is one of the inventors on patent US 10,349,899 B2 (Title: System and method for predicting hallucinations) held by the Swiss Federal Institute of Technology (EPFL) that covers a robotic system for the prediction of hallucinations for diagnostic purposes. OB, JP and FB are inventors on patent n° EP4407627A1 (Title: Numerosity estimation impairment measurement system) held by the Swiss Federal Institute of Technology (EPFL) that covers the implicit measure of presence hallucinations. The authors declare no further potential conflicts of interest with respect to the research, authorship, and/or publication of this article.

## Notes

### Author Declarations

The Cantonal Ethics Committee of Geneva gave ethical approval for this work (protocol reference 2019-02275), in accordance with the Declaration of Helsinki.

