## Supplementary information for "Neural Correlates of Cognitive Alterations and Minor and Structured Hallucinations in Parkinson’s Disease"

### Supplementary material

#### Supplementary methods

##### Neuropsychological testing

All patients completed the Parkinson's Disease Cognitive Rating Scale (PD-CRS)[1], a battery of neuropsychological tests assessing major cognitive domains such as executive functions, attention, visuospatial abilities, memory, and language. The PD-CRS allows the assessment of the frontal subcortical functions (frontal subcortical score) and of the temporal posterior cortical functions (posterior cortical score). As the posterior cortical score indicates more advanced cognitive decline and dementia, the current study focuses on the frontal subcortical score reflecting executive functioning and indicative of Mild Cognitive Impairment (MCI) if performances are low[1,2]. We tested subgroup differences in both posterior cortical and frontal subcortical PD-CRS scores with ANOVA.

##### Robot task inducing presence hallucinations

To induce PH-like states, or the feeling of presence, in laboratory settings, we employed a robotic system that was tested and used in previous work in healthy individuals and patients with PD[3–6]. By a somatomotor conflict, the robotic system can induce the sensation of someone nearby, and it is constituted by two parts: a commercial haptic interface (Phantom Omni, SensAble technologies) placed in front of a patient, and a custom-made three-degree-of-freedom robot behind the patient[5,7]. While performing the somatomotor experiment, patients were seated and blindfolded. They were also given headphones delivering constant white noise for an isolation from external distractions. Participants were instructed to move the front robot back and forth with a hand they felt most comfortable using. The back robot was reproducing these movements, applying tactile feedback on the participant's back with a pseudo-randomized delay of 0, 250 or 500 ms. In each trial, participants performed 10 poking movements, which were automatically counted, and subsequently answered "yes" or "no" to the question "Did you feel as if someone was standing close by — behind or next to you?". Each participant performed 12 trials per delay, that is, a total of 36 trials. There were breaks in between sessions to avoid physical discomfort or fatigue in patients.

We first examined the task responses with an ordinary least squares linear model, fitting the model to the proportion of "yes" answers in each delay in each individual. For our main data analysis, we first fitted a linear model to the proportion of "yes" answers in each delay in each individual. As based on previous work we expected that sensitivity to riPH should increase with an increasing delay [4,6], we then categorized individuals as either "responders" if the model's slope was larger than zero, or as "non-responders" otherwise. We first tested subgroup differences in the proportions of responders vs. non-responders by performing the Chi-square test in each subgroup and correcting for multiple comparisons with the false discovery rate (FDR) correction. For closer inspection, we also fitted a linear regression model to the proportions of "yes" answers in each delay to evaluate the effects of delay and subgroup.

##### Effects of nuisance variables

We tested whether there was a relationship between the identified LC and variables of no interest, such as age, years of education, duration of the disease, and the total LEDD, by Spearman's $\rho$ between the brain scores and the given variable in each patient subgroup. We then further examined the effect of the total LEDD by first regressing it out from the FC and then running the PLSC analysis as described above on the residualized data.

#### Supplementary results

##### Effects of nuisance variables

To test for potential effects of demographic and clinical variables that were not of interest, we evaluated correlations of the brain scores with age, education, disease duration, disease stage, and medication dose within each patient subgroup. We found no significant relationships with the nuisance variables, except in the total LEDD that was significantly correlated with brain scores in the MH subgroup ($\rho$ = -0.659, $p_{FDR}$ = 0.009) (**Supplementary Fig. 6**). Notably, there was a marginally significant association between the total LEDD and frontal subcortical scores in the MH subgroup ($\rho$ = -0.546, $p$ = 0.019, $p_{FDR}$ = 0.058) (**Supplementary Fig. 7a**). Therefore, as expected, in a follow-up PLSC analysis in which FC matrices were residualized with respect to the total LEDD the association between brain scores and frontal subcortical scores in the MH subgroup was attenuated (**Supplementary Fig. 7b**). Importantly, dopamine agonists, that have been previously associated with hallucinations in PD[8], were not correlated with the brain scores, nor did they significantly differ between the patient subgroups. Further investigations are needed to fully understand the relationship between the total LEDD and frontal subcortical scores of the PD-CRS.

#### Supplementary figures

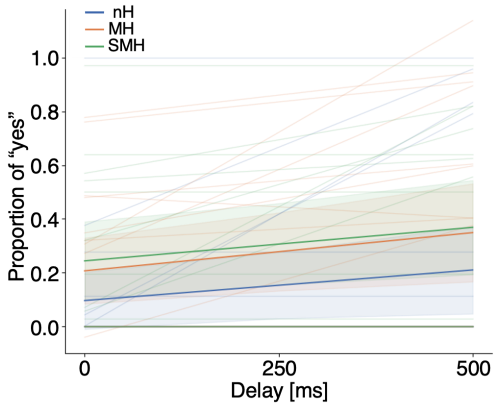


**Supplementary Figure 1. Robot task responses in stimulation delays.** The plot shows individual regression lines fitted to the proportion of "yes" in each delay as the lighter lines and the mean in patient subgroups as the darker lines with the shaded area representing the 95 % confidence interval.

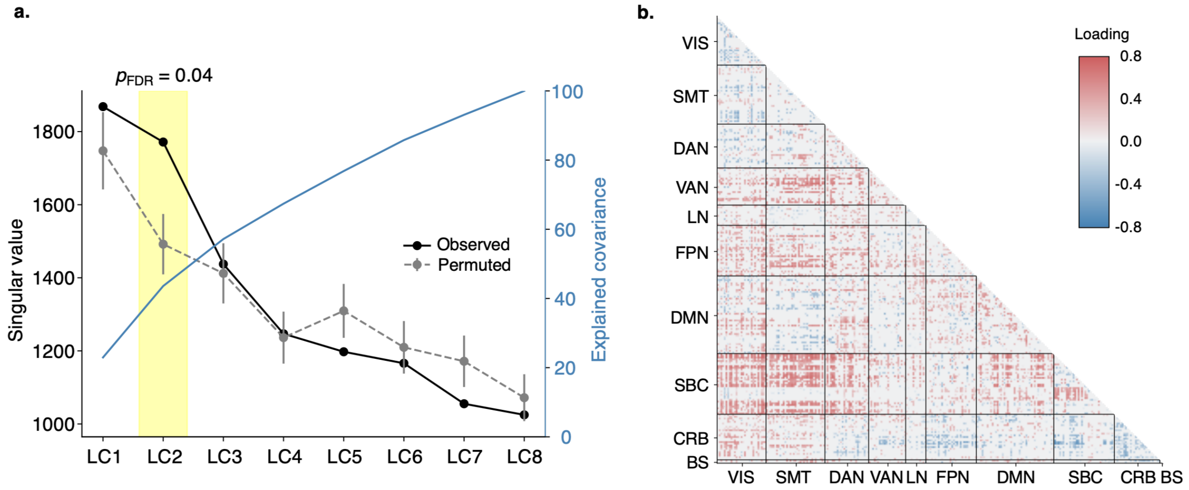


**Supplementary Figure 2. Evaluation of LC significance in the PLSC analysis and significant FC in LC2.** **a.** We tested eight LCs against the chance level obtained by a permutation test. The second LC was significant after FDR. **b.** The matrix shows loadings of all significant edges with BSR > 2 as suggested by Krishnan et al., 2011.

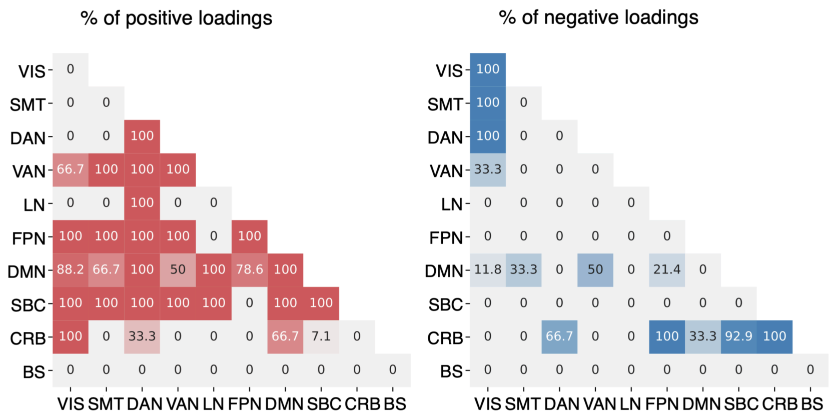


**Supplementary Figure 3. Proportion of positive and negative loadings in networks.**

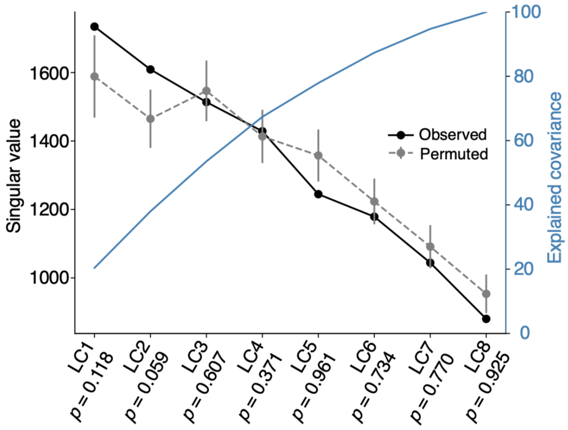


**Supplementary Figure 4. The PLSC analysis using the posterior cortical scores of PD-CRS.** As a control analysis, we also ran the PLSC analysis with the posterior cortical scores of the PD-CRS instead of the frontal subcortical scores. The analysis did not reveal any significant LC.

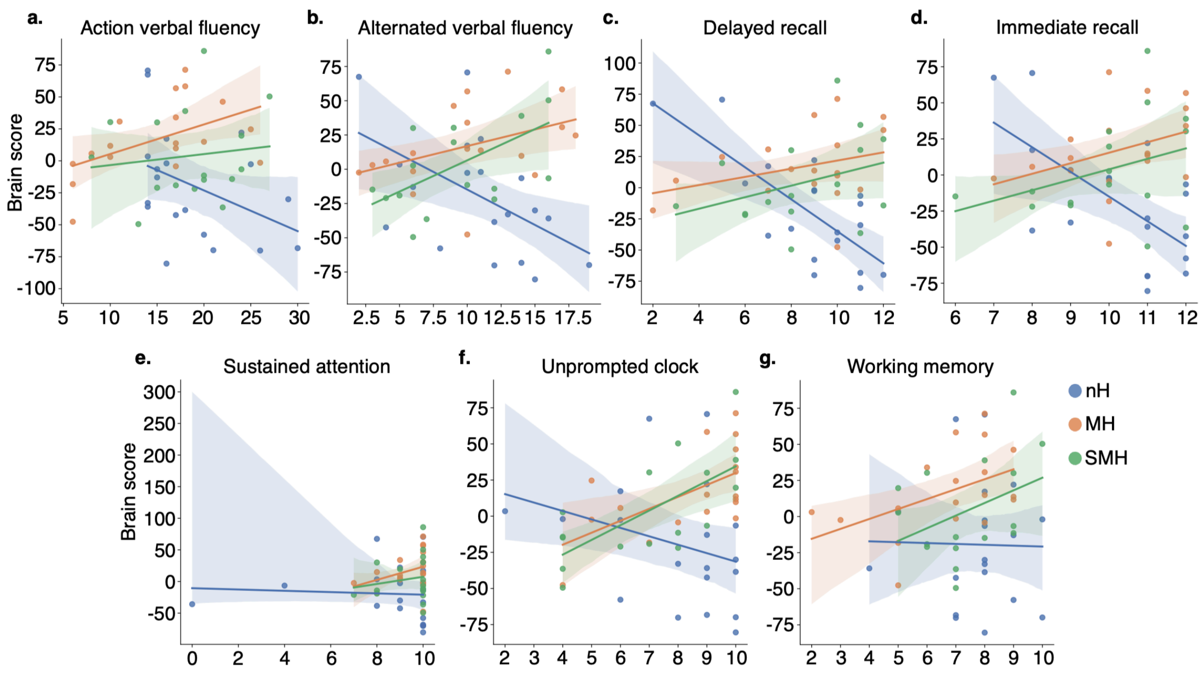


**Supplementary Figure 5. Relationship between brain scores and subtests of the frontal subcortical score of the PD-CRS.** The regression plots depict the relationship between the brain and **a.** action verbal fluency, where the MH subgroup was significant ($\rho = 0.57$, $p_{FDR} = 0.041$), **b.** alternated verbal fluency, in which significant correlation was found in nH ($\rho = -0.511$, $p_{FDR} = 0.045$), and SMH ($\rho = 0.542$, $p_{FDR} = 0.045$), **c.** delayed recall, where nH was significant ($\rho = -0.612$, $p_{FDR} = 0.016$), **d.** immediate recall, in which also nH was significant ($\rho = -0.548$, $p_{FDR} = 0.045$), **e.** sustained attention, **f.** unprompted clock, where we found significance in MH ($\rho = 0.567$, $p_{FDR} = 0.021$) and SMH ($\rho = 0.657$, $p_{FDR} = 0.017$), and **g.** working memory subscore.

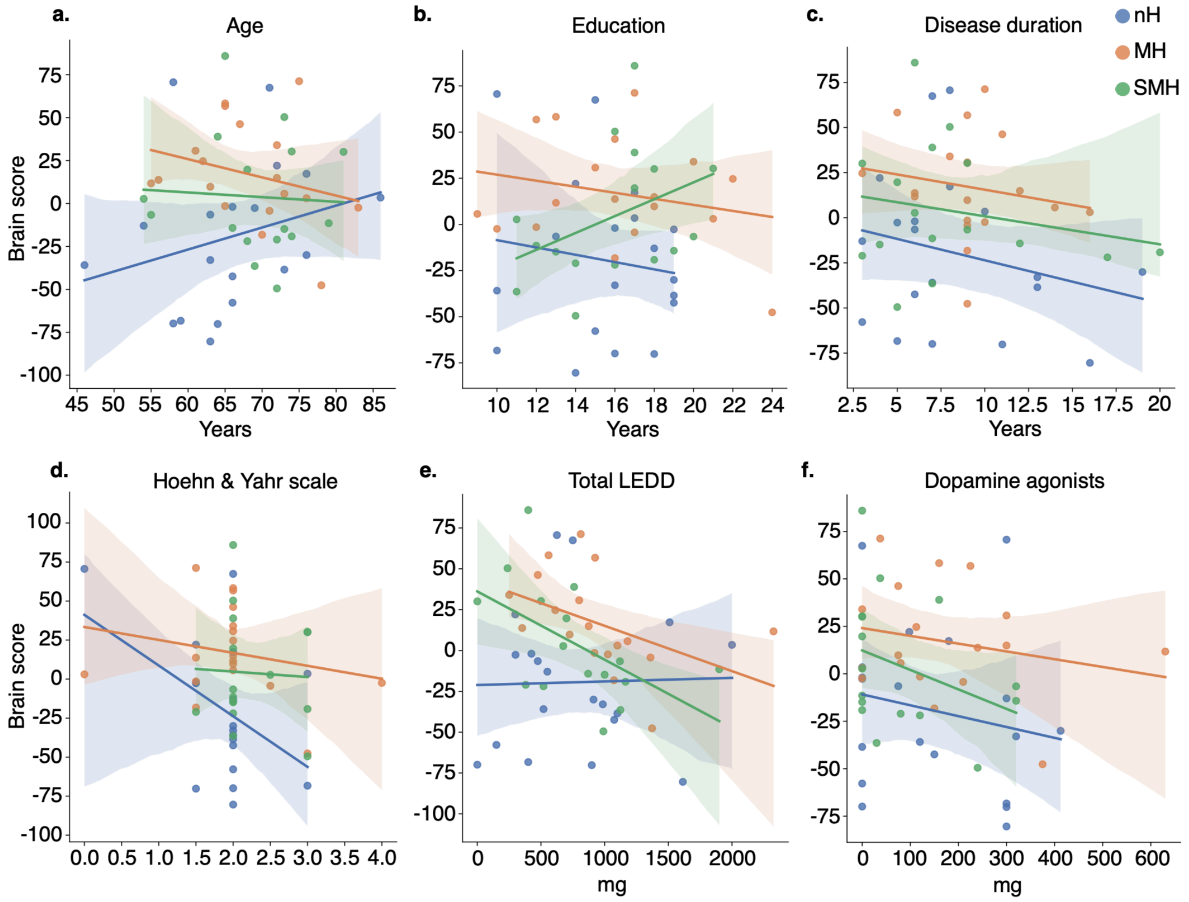


**Supplementary Figure 6. Effects of nuisance variables.** Variables including **a.** age, **b.** education, **c.** disease duration, **d.** disease progression measured by the Hoehn-Yahr scale, and **f.** dose of dopamine agonists were not significantly correlated with the brain score in any group. The only significant correlation was found in **e.** the total LEDD in the MH subgroup ($\rho$ = -0.659, $p_{FDR}$ = 0.009), and in the SMH subgroup before FDR correction ($\rho$ = -0.5, $p$ = 0.049, $p_{FDR}$ = 0.073).

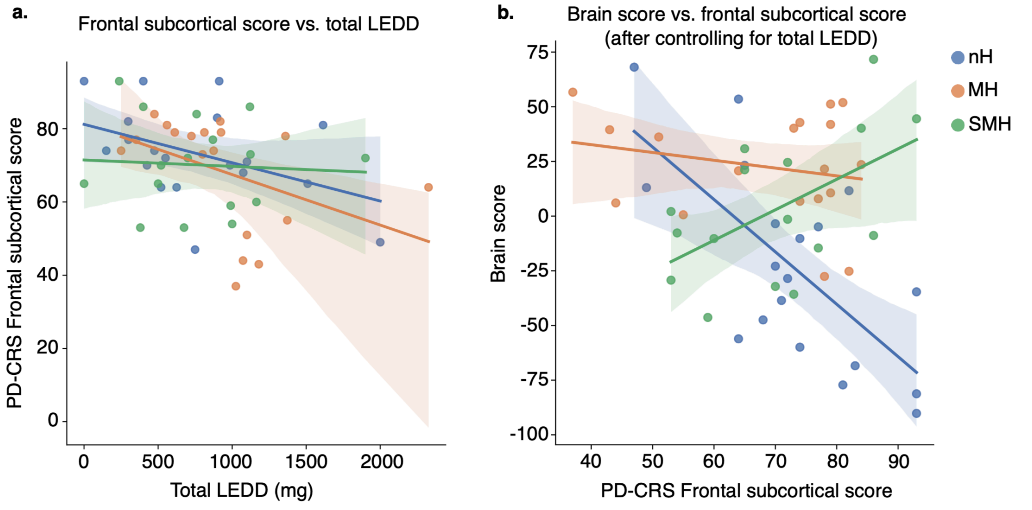


**Supplementary Figure 7. Effects of the total LEDD. a.** There was a marginally significant relationship between the total LEDD and the frontal subcortical score of PD-CRS in the mH subgroup ($\rho$ = -0.546, $p$ = 0.019, $p_{FDR}$ = 0.058**). b.** After regressing out the effects of the total LEDD from the FC entering the PLSC analysis the relationship between the brain scores and the frontal subcortical scores of the PD-CRS in the mH subgroup was no longer significant.
